# Reducing false reassurance following negative results from asymptomatic coronavirus (Covid-19) testing: an online experiment

**DOI:** 10.1101/2021.08.03.21261482

**Authors:** Eleonore Batteux, Stefanie Bonfield, Leah Ffion Jones, Holly Carter, Natalie Gold, Richard Amlôt, Theresa M. Marteau, Dale Weston

## Abstract

**Objectives:** Individuals who receive a negative lateral flow coronavirus (Covid-19) test result may misunderstand it as meaning ‘no risk of infectiousness’, giving false reassurance. This experiment tested the impact of adding information to negative test result messages about (a) residual risk and (b) need to continue protective behaviours.

**Design:** 4 (residual risk) × 2 (post-test result behaviours) between-subjects design.

**Setting:** Online.

**Participants:** 1200 adults from a representative UK sample recruited via Prolific (12-15 March 2021).

**Interventions:** Participants were randomly allocated to one of eight messages. Residual risk messages were: 1) ‘Your coronavirus test result is negative’ (control); 2) Message 1 plus ‘It’s likely you were not infectious when the test was done’ (Current NHS Test & Trace); 3) Message 2 plus ‘But there is still a chance you may be infectious’ (Elaborated NHS T&T); 4) Message 3 plus infographic depicting residual risk (Elaborated NHS T&T + infographic). Each message contained either no additional information or information about behaviour, i.e. the need to continue following guidelines and protective behaviours.

**Outcome measures:** (i) proportion understanding residual risk of infectiousness and (ii) likelihood of engaging in protective behaviours (score range 0-7).

**Results:** The control message decreased understanding relative to the current NHS T&T message: 54% vs 71% (AOR=0.37 95% CI [0.22, 0.61], *p*<.001). Understanding increased with the elaborated NHS T&T (89%; AOR=3.27 95% CI [1.78, 6.02], *p*<.001) and elaborated NHS T&T + infographic (91%; AOR=4.03 95% CI [2.14, 7.58], *p*<.001) compared to current NHS T&T message. Likelihood of engaging in protective behaviours was unaffected by information (*F*(1,1192)=0.43, *p*=.513), being high (*M*=6.4, *SD*=0.9) across the sample.

**Conclusions:** The addition of a single sentence (‘But there is still a chance you may be infectious’) to current NHS Test & Trace wording increased understanding of the residual risk of infection.

**Trial registration:** Open Science Framework: https://osf.io/byfz3/

## Introduction

As part of the global effort to reduce the transmission of coronavirus (Covid-19), asymptomatic testing via rapid antigen tests such as lateral flow devices (LFDs) has become widespread^1^. LFDs have high specificity (over 99%), meaning they are highly likely to correctly identify people who are not infectious^2^. However, they have lower sensitivity and can incorrectly provide a negative test result in up to 50% of asymptomatic positive Covid-19 cases^2^, either due to lower viral load^3^ or improper sampling techniques, which are more likely when tests are conducted unsupervised^3^. This means individuals could be told they are not infectious when in fact they are. Given this, individuals who receive a negative test result (i.e. the majority) need to understand the residual risk of infectiousness and the need to continue following government guidelines.

The extent to which people understand the residual risk of infection after a negative asymptomatic Covid-19 test result is not known. Research on negative test results in cancer screening suggests that just 52% of people have a correct understanding of residual risk^4^. This can produce false reassurance and detrimental changes to behaviour^5^, where individuals may be less concerned if they experience symptoms of an infection or disease in the future or may reduce their engagement in protective behaviours^6-9^. This is akin to the ‘health certificate effect’ whereby a negative result can reduce motivation to protect oneself against a health threat^6^. In the context of Covid-19, if people take a negative test result to mean no risk of infection, this could lead to reduced adherence to Covid-19 guidelines^10^.

Importantly, the way in which negative test results are communicated can affect understanding and behaviour. For example, communicating that there is still a risk of cervical cancer after a negative screening result increases understanding compared to communicating that the residual risk is lower than for the average person (OR 5.46)^5^. In the context of Covid-19, communicating residual risk with a negative PCR test result makes people more likely to agree that a symptomatic individual should continue to self-isolate, compared to not communicating it (96% vs 83)^11^. Furthermore, graphical representations of risk have been found to increase understanding in healthcare contexts^12,13^. For example, the addition of an icon array to numerical risk information can improve the accuracy of risk estimates in medical scenarios (medium effect size)^12^. This shows that emphasising residual risk in negative test results both visually and verbally could increase understanding that a risk remains.

Test result messages also offer an opportunity to communicate the need to continue adhering to protective behaviours after a negative result, which might not be immediately clear if individuals are given a negative result but told that they could still be infectious. Unambiguous behavioural instructions and guidelines in Covid-19 messaging are encouraged by The British Psychological Society^14^ and can provide the knowledge and capability people need to engage in protective behaviours^15^. It is also likely to be valuable given that responses to a health threat are influenced by whether an individual believes there are behaviours they can engage in to reduce or alleviate the risk^16^.

At the time of writing, the NHS Test and Trace (T&T) negative result messaging communicates some residual risk which is positively framed (see Box 1). However, perceptions of risk or uncertainty have been shown to increase when messages contain negative framing or if positive and negative framing are combined, compared to positive framing alone^17-19^. The addition of a negatively framed sentence to the existing NHS T&T messaging could therefore improve understanding. Post-test result behaviours are also included in existing messaging^20^, but to our knowledge have not been evaluated.

#### Box 1: Residual risk and post-test result behaviours

##### Residual risk messages

###### No residual risk information

‘Your coronavirus test result is negative.’

###### Current NHS Test & Trace

‘Your coronavirus test result is negative. It’s likely you were not infectious when the test was done.’

###### Elaborated NHS Test & Trace

‘Your coronavirus test result is negative. It’s likely you were not infectious when the test was done. But there is still a chance you may be infectious.’

###### Elaborated NHS Test & Trace + infographic

*‘Your coronavirus test result is negative. It’s likely you were not infectious when the test was done. But there is still a chance you may be infectious.’* + infographic (see Supplementary Material)

##### Post-test result behaviours

This means you should continue to follow all government guidance to reduce transmission of the virus. You must stay at home. You must not leave or be outside of your home except where necessary.

Remember - ‘Hands. Face. Space.’

- hands – wash your hands regularly and for at least 20 seconds
- face – wear a face covering in indoor settings where social distancing may be difficult, and where you will come into contact with people you do not normally meet
- space – stay 2 metres apart from people you do not live with where possible, or 1 metre with extra precautions in place (such as wearing face coverings)

Given the dearth of research examining the understanding of residual risk and behaviours following a negative Covid-19 LFD test, we conducted an online experiment examining the impact of communicating about residual risk and protective behaviours following a negative test result. The protocol was preregistered on Open Science Framework (OSF) (https://osf.io/byfz3/) and hypotheses were as follows:

### Hypothesis 1

Understanding of residual risk is (a) increased by adding existing NHS T&T messaging compared to no information about residual risk (control) and (b) increased further by adding an elaborated message and an infographic.

### Hypothesis 2

Expectations to follow coronavirus guidelines are higher when messages contain information about the need for continued engagement in protective behaviours.

## Method

### Design

Participants were randomly allocated to a message in a 4 (residual risk) ⨯ 2 (post-test result behaviours) between-subjects design (see Box 1).

### Participants

A cross-stratified quota sample of 1207 UK adults representative of the UK population based on sex, age and ethnicity was recruited via the online platform Prolific (https://www.prolific.co/) between 12-15 March 2021, during the third national lockdown in England. A quota sample fills predetermined targets so that demographic characteristics are representative of the general population. Participants are prevented from completing the experiment if they belong to a quota that has already been filled.

### Power

The power analyses conducted with G*Power (version 3.1) indicated that a sample of 1095 was needed to test the hypotheses. For Hypothesis 1, given the lack of prior data, a power analysis for a logistic regression could not be conducted and was based on a chi-square test instead. A sample of 547 can detect a difference between two groups with a small effect size (*w*=0.12), using a chi-square test with α=0.05 and power >.80. For 4 groups, it was estimated that double the sample size was needed, i.e. 1094 participants. For Hypothesis 2, 1095 participants can detect a small effect size (*f*=0.10) using a between-subjects ANOVA with α=0.05 and power >.80. We planned to exclude participants who failed an attention check (see Supplementary Material). As 10% of participants were expected to fail it, 1205 participants were needed to ensure 1095 participants could be included in the analysis.

### Messages

Participants imagined they had taken a lateral flow test and received one of 8 messages (see Box 1 and Supplementary Material). The messages incrementally varied the level of residual risk communicated. The control condition provided no information about residual risk, the current NHS T&T^1^ condition adds positively framed information about residual risk to the control message, the elaborated NHS T&T condition adds negatively framed information about residual risk to the existing NHS T&T messaging, and the elaborated NHS T&T and infographic condition adds an infographic with numerical residual risk information to the elaborated NHS T&T message. The infographic is based on 1% prevalence, 99% specificity and 50% sensitivity and includes a) a flow chart illustrating among a given population the number of positive and negative test results within individuals who are infected and those who are not and b) an icon array demonstrating the proportion of those receiving a negative result who are actually infected.

The message also contained either none or some information about the need to maintain adherence to protective behaviours following a negative test result, as listed on UK government guidance under national lockdown in March 2021^21^. This information indicates that people should continue to follow all government guidance and reminds them of key protective behaviours (hands, face, space).

### Primary outcome measures

Primary outcome measures were understanding of residual risk and behavioural expectations to follow Covid-19 guidelines (see Supplementary Material). Understanding of residual risk was measured by asking participants to identify the correct statement from four options: ‘I am not infectious with coronavirus’, ‘I am most likely not infectious with coronavirus’ (correct), ‘I am most likely infectious with coronavirus’, ‘I am infectious with coronavirus’.

Behavioural expectations to follow Covid-19 guidelines were measured with specific protective behaviour questions and a general question. Six protective behaviours were measured with a 7-point scale question: ‘After receiving this test result, how likely is it that you would engage in the following behaviours because of coronavirus?’ (behaviours: social distancing, hand washing, wearing a face covering, avoiding meeting others, working from home, avoiding public transport; 1-very unlikely to 7-very likely), taken from a previous study^22^. There was good reliability between questions (Cronbach’s α = .86) which were averaged to provide an overall score of behavioural expectation. The general question was adapted from previous studies^22,23^: ‘Having received this test result, how strictly would you follow coronavirus guidelines now compared to before taking the test?’ (1-a lot less strictly; 7-a lot more strictly).

### Secondary outcome measures

Secondary outcome measures were confidence in understanding, perceived test accuracy and testing uptake expectations (see Supplementary File). Participants were asked how confident they were in their understanding of residual risk (1-not at all confident; 5-extremely confident). They were asked how accurate they thought rapid lateral flow tests were (1-very inaccurate; 7-very accurate) and how likely they were to take a rapid lateral flow test in the future (1-very unlikely; 7-very likely) as there is a risk that communicating residual risk could give the impression that antigen tests are inaccurate and not worth taking.

### Other measures

Participants were asked about their previous testing experience, including the last time they took a coronavirus test and what type of test it was (see Supplementary Material). A frequently used numeracy question was administered to assess their understanding of proportions^24^. Those who received the message containing the infographic were asked how easy it was to understand (1-very difficult; 5-very easy) and any suggestions for improvements (text box). An attention check (a multiple choice question asking participants not to select an option) and a recognition question (asking participants to select the test result they received) were included to evaluate participant attention throughout the study. Finally, participants were asked demographic questions (gender, age, ethnicity, UK region, highest level of education).

### Procedure

Participants were recruited via Prolific and then directed to the study on Qualtrics. They were asked to imagine they had taken a lateral flow test as part of a local mass asymptomatic testing programme, similar to those taking place in the UK^25^. They then received a message about the result of their test, to which they were randomised using the Qualtrics randomisation function, and answered a series of questions (see Supplementary Material). Participants were unaware of the condition they were allocated to and paid at a rate of £25 per hour (i.e. £2.10 for a 5-minute experiment).

### Patient and public involvement

Patients and/or the public were not involved in the development of the study due to the rapid nature of this research. However, the experiment was piloted with 16 participants to ensure it ran smoothly and that there were no errors. Those who took part in the pilot were able to provide feedback to researchers on the study.

### Analysis

Pre-registered analyses were conducted using SPSS (version 27) with a significance level of *p*<.05. To test Hypothesis 1, a binomial logistic regression was conducted with residual risk, post-test result behaviour and an interaction term as predictors of understanding (coded as correct: ‘I am most likely not infectious with coronavirus’, or incorrect: all other responses). Group 2 (current NHS T&T) was used as the reference category for the residual risk predictor. Age, gender, ethnicity, education, location and numeracy were added to the model as covariates. To test hypothesis 2, a 4 (residual risk) ⨯ 2 (post-test result behaviour) between-subjects ANOVA was conducted on specific protective behaviours. Expected engagement in specific behaviours was negatively skewed, which was corrected with pre-planned logarithmic transformation. Other analyses reported are exploratory.

## Results

Of the 1207 participants who completed the study, 7 (0.6%) failed the attention check and were excluded from the analysis. A breakdown of the demographic characteristics of the remaining 1200 participants can be found in Table 1.

**Table 1:** Participant demographic characteristics

| Demographic characteristic | n | % |
| --- | --- | --- |
| *Gender* |  |  |
| Male | 582 | 48.5% |
| Female | 615 | 51.2% |
| Non-binary | 1 | 0.1% |
| Prefer not to say | 2 | 0.2% |
| *Age* |  |  |
| 18-24 | 127 | 10.6% |
| 25-64 | 902 | 75.2% |
| 65+ | 171 | 14.2% |
| *Education* |  |  |
| GCSE or equivalent | 221 | 18.4% |
| A level or equivalent | 298 | 24.8% |
| Undergraduate degree | 482 | 40.2% |
| Postgraduate degree | 199 | 16.6% |
| *Ethnicity* |  |  |
| White - British | 906 | 75.5% |
| White - Other | 113 | 9.4% |
| Asian | 98 | 8.2% |
| Black | 41 | 3.4% |
| Mixed | 32 | 2.7% |
| Other | 10 | 0.9% |
| *UK region* |  |  |
| NI/Scotland/Wales | 162 | 13.4% |
| England – South | 316 | 26.3% |
| England – London | 155 | 12.9% |
| England – Midlands | 268 | 22.3% |
| England – North | 299 | 24.9% |
| *Testing experience* |  |  |
| Yes – PCR | 235 | 19.6% |
| Yes – LFT | 281 | 23.4% |
| Yes – Other (e.g. antibody) | 33 | 2.8% |
| Yes – Don't know | 44 | 3.7% |
| None | 607 | 50.6% |

### Understanding of residual risk

Understanding varied by residual risk message as outlined in Hypothesis 1 (see Table 2), as shown by a binomial logistic regression in Table 3. Those who saw the existing NHS T&T message were more likely to have a correct understanding of residual risk (71.1%) than those in the control group who received no information about residual risk (54.3%) (AOR=0.58 95% CI [0.35, 0.97], χ^2^(1)=4.32, *p=*.038) (see Figure 1). Those who saw the elaborated NHS T&T message were more likely to have a correct understanding (88.7%) than those who saw the existing NHS T&T message (AOR=3.31 95% CI [1.68, 6.51], χ^2^(1)=11.95, *p*<.001). This was also the case for the elaborated NHS T&T message with the infographic (90.7%) (AOR=5.31 95% CI [2.54, 11.10], χ^2^(1)=19.73, *p*<.001). However, understanding in this condition was not significantly higher than the elaborated NHS T&T message alone (χ^2^(1)=0.60, *p*=.437). Understanding was lower among those with lower education, those with lower numeracy and those from Black and Mixed ethnicity compared to White British ethnicity (see Table 3). The model correctly classified 78.9% of cases and was a good fit to the data according to the Hosmer–Lemeshow test (χ^2^ (8)=4.77, p=.782).

**Table 2:** Primary and secondary outcomes (%(n); mean (SD)) by experimental group

|  | Residual risk |  |  |  | Post-test result behaviours |  |
| --- | --- | --- | --- | --- | --- | --- |
|  | Control<br>(n=300) | NHS T&T<br>(n=298) | Elaborated<br>(n=302) | Infographic<br>(n=300) | None<br>(n=602) | Included<br>(n=598) |
| **Primary measures** |  |  |  |  |  |  |
| **Understanding** |  |  |  |  |  |  |
| I am not infectious | 45.3%<br>(n=136) | 28.2%<br>(n=84) | 9.6%<br>(n=29) | 7.7% (n=23) | 19.6%<br>(n=118) | 25.8%<br>(n=154) |
| I am most likely not infectious* | 54.3%<br>(n=163) | 71.1%<br>(n=212) | 88.7%<br>(n=268) | 90.7%<br>(n=272) | 79.7%<br>(n=480) | 72.7%<br>(n=435) |
| I am most likely infectious | 0% (n=0) | 0.3% (n=1) | 1.3% (n=4) | 0.7% (n=2) | 0.5% (n=3) | 0.7% (n=4) |
| I am infectious | 0.3% (n=1) | 0.3% (n=1) | 0.3%<br>(n=1) | 1.0% (n=3) | 0.2% (n=1) | 0.8% (n=5) |
| **Specific behaviours** |  |  |  |  |  |  |
| Average | 6.40 (0.9) | 6.46 (0.8) | 6.42 (0.9) | 6.33 (1.1) | 6.39 (0.9) | 6.41 (0.9) |
| Social distancing | 6.52 (1.0) | 6.55 (1.0) | 6.53 (1.0) | 6.46 (1.2) | 6.53 (1.0) | 6.50 (1.1) |
| Hand washing | 6.45 (1.0) | 6.50 (1.0) | 6.46 (1.1) | 6.41 (1.2) | 6.48 (1.1) | 6.44 (1.1) |
| Face covering | 6.70 (0.8) | 6.71 (0.9) | 6.71 (0.9) | 6.55 (1.3) | 6.70 (0.9) | 6.63 (1.1) |
| Avoid meeting others | 6.20 (1.3) | 6.21 (1.3) | 6.15 (1.3) | 6.00 (1.5) | 6.09 (1.3) | 6.18 (1.3) |
| Work from home | 6.19 (1.5) | 6.32 (1.4) | 6.24 (1.4) | 6.21 (1.4) | 6.20 (1.5) | 6.28 (1.4) |
| Avoid public transport | 6.28 (1.4) | 6.47 (1.2) | 6.44 (1.2) | 6.34 (1.3) | 6.35 (1.3) | 6.43 (1.2) |
| **Secondary measures** |  |  |  |  |  |  |
| Expectations to follow guidelines | 4.23 (0.9) | 4.18 (0.8) | 4.25 (0.9) | 4.32 (0.9) | 4.19 (0.8) | 4.30 (0.8) |
| Confidence in understanding | 4.17 (0.8) | 4.35 (0.8) | 4.23 (0.8) | 4.32 (0.8) | 4.24 (0.8) | 4.29 (0.8) |
| Perceived testing accuracy | 5.71 (1.1) | 5.71 (1.1) | 5.61 (1.1) | 5.95 (1.0) | 5.75 (1.1) | 5.74 (1.1) |
| Future testing expectations | 5.90 (1.6) | 5.92 (1.6) | 5.88 (1.6) | 5.99 (1.6) | 5.90 (1.6) | 5.95 (1.6) |

Notes: \* refers to a correct understanding of residual risk. Confidence is on a 5-point scale and other continuous variables on a 7-point scale.

**Table 3:** Logistic regression predicting correct understanding of residual risk.

|  | AOR | 95% CI | Wald | p |
| --- | --- | --- | --- | --- |
| Intercept | 1.26 | 0.49, 3.19 | 0.23 | .633 |
| *Residual risk* |  |  |  |  |
| Control | 0.58 | 0.35, 0.97 | 4.32 | **.038** |
| NHS T&T (reference) |  |  |  |  |
| Elaborated T&T | 3.31 | 1.68, 6.51 | 11.95 | **<.001** |
| Elaborated T&T + infographic | 5.31 | 2.54, 11.10 | 19.73 | **<.001** |
| *Post-test result behaviours* |  |  |  |  |
| Without (reference) |  |  |  |  |
| With | 0.82 | 0.49, 1.38 | 0.58 | .447 |
| *Residual risk \* Post-test result behaviours* |  |  |  |  |
| NHS T&T * With (reference) |  |  |  |  |
| Control * With | 0.64 | 0.31, 1.31 | 1.50 | .220 |
| Elaborated T&T * With | 0.95 | 0.38, 2.38 | 0.01 | .919 |
| Elaborated T&T + infographic * With | 0.73 | 0.28, 1.92 | 0.40 | .525 |
| *Gender<sup>2</sup>* |  |  |  |  |
| Male (reference) |  |  |  |  |
| Female | 1.06 | 0.79, 1.43 | 0.15 | .698 |
| *Age* | 0.99 | 0.98, 1.00 | 1.62 | .204 |
| *Education* |  |  |  |  |
| GCSE or equivalent (reference) |  |  |  |  |
| A-level or equivalent | 1.84 | 1.20, 2.83 | 7.72 | **.005** |
| Undergraduate | 2.70 | 1.80, 4.06 | 23.01 | **<.001** |
| Postgraduate | 4.83 | 2.79, 8.34 | 31.55 | **<.001** |
| *Ethnicity* |  |  |  |  |
| White British (reference) |  |  |  |  |
| White Other | 0.79 | 0.46, 1.38 | 0.68 | .411 |
| Asian | 0.60 | 0.34, 1.07 | 2.98 | .084 |
| Black | 0.34 | 0.16, 0.74 | 7.45 | **.006** |
| Mixed | 0.36 | 0.15, 0.90 | 4.77 | **.029** |
| Other | 0.66 | 0.12, 3.60 | 0.23 | .633 |
| *Location* |  |  |  |  |
| London (reference) |  |  |  |  |
| Northern Ireland | 1.09 | 0.27, 4.46 | 0.01 | .907 |
| Scotland | 0.81 | 0.41, 1.60 | 0.37 | .541 |
| Wales | 0.64 | 0.29, 1.42 | 1.21 | .271 |
| South England | 1.10 | 0.64, 1.86 | 0.11 | .738 |
| Midlands | 1.47 | 0.85, 2.56 | 1.89 | .169 |
| North England | 0.87 | 0.51, 1.47 | 0.23 | .596 |
| *Numeracy* |  |  |  |  |
| Incorrect (reference) |  |  |  |  |
| Correct | 1.67 | 1.12, 2.42 | 7.49 | **.006** |

**Figure 1:**
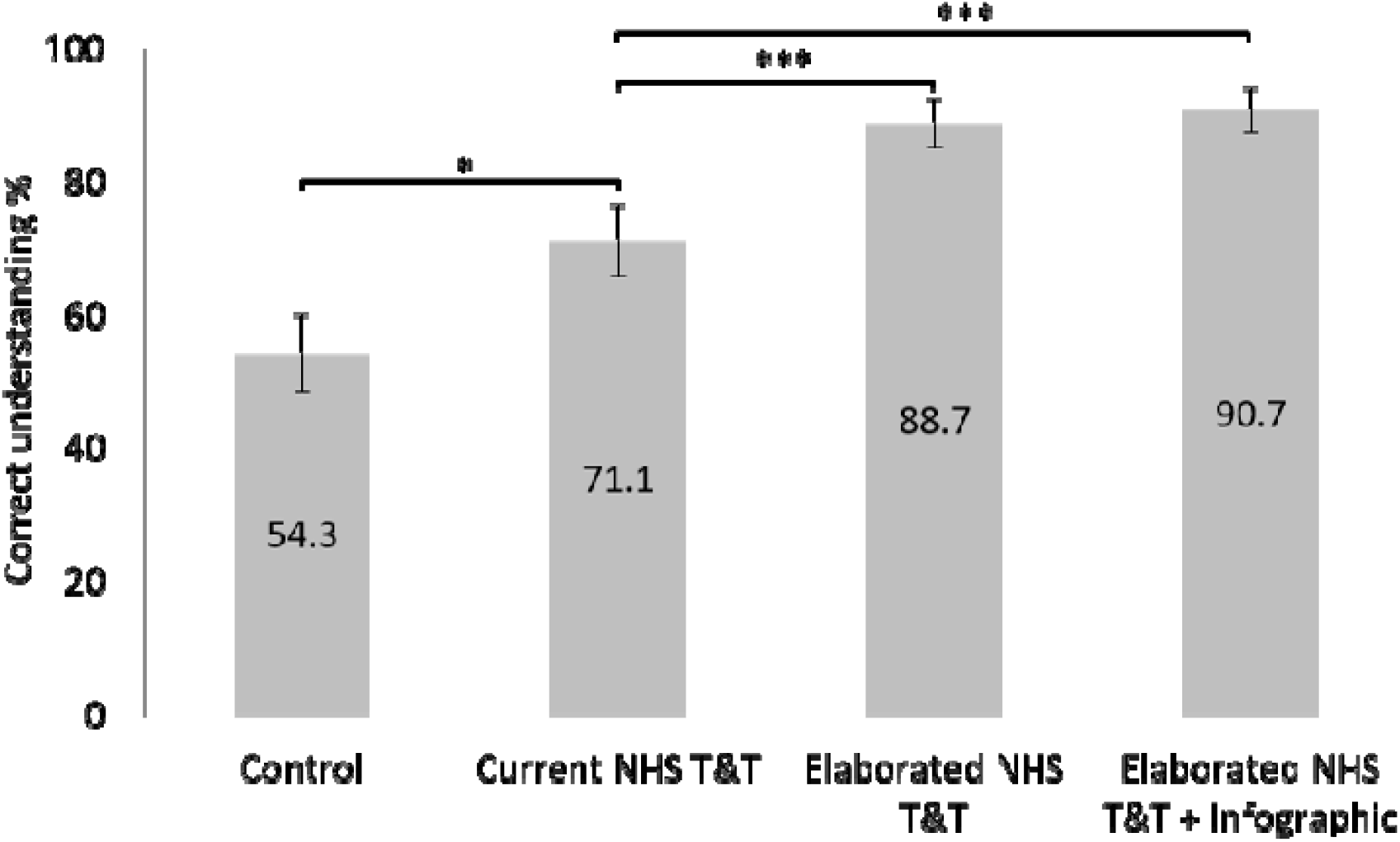
Percentage of participants with a correct understanding of residual risk by residual risk experimental group. Error bars represent 95% confidence intervals. Significance levels are based on the logistic regression in Table 3). * refers to p<.05, ** refers to p<.01, *** refers to p<.001.

### Confidence in understanding

As planned, we explored whether residual risk messages affected confidence in understanding among those who were correct (76.3%), to assess the effectiveness of messages beyond understanding. Residual risk information affected confidence (F(3,907)=10.94, p<.001, η2=.04), with the control group being less confident (M=3.93, SD=0.77) than existing NHS T&T (M=4.36, SD=0.73, p<.001), elaborated NHS T&T (M=4.24, SD=0.81, p<.001) and elaborated NHS T&T with the infographic (M=4.32, SD=0.80, p<.001) according to post-hoc tests (Tukey). There were no significant differences between other groups. Neither post-test result behaviours (F(1,907)=1.06, p=.304, η2<.01) nor the interaction between residual risk and post-test result behaviours (F(3,907)=0.53, p=.664, η2<.01) had a significant effect on confidence.

### Post-test result behaviours

Communicating about the need to maintain protective behaviours following a negative test result did not significantly increase expected engagement in protective behaviours (*F*(1,1192)=0.38, *p*=.536, η2<.01), which does not support Hypothesis 2. Neither residual risk (*F*(3,1192)=0.83, *p*=.476, η2<.01) nor the interaction between residual risk and post-test result behaviours (*F*(3,1192)=0.66, *p*=.579, η2<.01) had a significant effect on expected engagement in protective behaviours.

Those who received information on the need to maintain protective behaviours had higher expectations that they would follow guidelines (*M*=4.30, *SD*=0.80) than those who did not (*M*=4.19, *SD*=0.80) (*F*(1,1192)=5.26, *p*=.022, η2=.004), in line with Hypothesis 2. Neither residual risk (*F*(3,1192)=1.56, *p*=.199, η2<.01) nor the interaction between residual risk and post-test result behaviours (*F*(3,1192)=0.56, *p*=.644, η2<.01) had a significant effect on expectations of following coronavirus guidelines.

### Perceived accuracy

Perceived accuracy of lateral flow tests (see Table 2) was influenced by residual risk condition (*F*(3,1192)=5.38, *p*=.001, η2=.01). Those who saw the infographic perceived lateral flow tests as more accurate (*M*=5.95, *SD*=1.00) than those who saw no residual risk information (*M*=5.71, *SD*=1.10; *p*=.034), existing NHS T&T messaging (*M*=5.71, *SD*=1.10; *p*=.029) and elaborated NHS T&T messaging (*M*=5.61, *SD*=1.1; *p*=.001) according to post-hoc tests (Tukey). There were no significant differences between other groups. Neither post-test result behaviours (*F*(1,1192)=0.06, *p*=.809, η2<.01) nor their interaction with residual risk (*F*(3,1192)=0.45, *p*=.714, η2<.01) affected perceived accuracy.

### Uptake expectations

Expectations to engage in asymptomatic lateral flow testing in the future (see Table 2) were not affected by residual risk information (*F*(3,1192)=0.27, *p*=.849, η2<.01), posttest result behaviours (*F*(1,1192)=0.37, *p*=.545, η2<.01) or their interaction (*F*(3,1992)=1.30, *p*=.272, η2<.01).

### Association between understanding and behavioural expectations

We explored whether those who had a correct understanding (n=915) were more likely to engage in protective behaviours compared to those who reported that there was no residual risk (n=272), bearing in mind participants were not randomised to each group. Those with a correct understanding did not have higher expected engagement in protective behaviours (*M*=6.40, *SD*=0.95) than those who believed there was no residual risk (*M*=6.38, *SD*=0.87) (*t*=0.47, df=1185, *p*=.641). Those with a correct understanding had lower expectations that they would follow guidelines (*M*=4.19, *SD*=0.73) than those who believed there was no residual risk (*M*=4.35, *SD*=1.07) (*t*=2.24, df=349.37, *p*=.026).

## Discussion

Enhanced communication of residual risk information in negative asymptomatic coronavirus test results improved understanding of residual risk, without evidence that it decreased the perceived accuracy of LFDs or testing uptake expectations. The elaborated NHS T&T message was better understood than the current NHS T&T message (89% vs 71% correct), which itself was more effective than giving no residual risk information (54% correct). The elaborated NHS T&T message added residual risk information which was negatively framed (‘But there is still a chance you may be infectious’) to the current NHS T&T message, which was positively framed (‘It’s likely you were not infectious when the test was done’). This study therefore echoes previous findings on negatively framed communications of residual risk^5^,to which it adds that combining positive and negative framing is also effective, as has been found in other health contexts^19^.

Adding an infographic with an icon array of residual risk did not significantly improve understanding relative to the elaborated NHS T&T message. This may be due to a ceiling effect given that the elaborated NHS T&T message increased understanding to nearly 90%. Although it contrasts with previous findings on the effectiveness of infographics^12, 13^, there is a precedent for them not increasing understanding of residual risk relative to verbal communications^4^. The infographic increased perceptions of testing accuracy, which could be because it includes numerical information which participants associated with accuracy. Indeed, this seems akin to the ‘seductive allure effect’ whereby people find psychological explanations more convincing when presented alongside irrelevant neuroscience information^26^. Furthermore, this did not result in differences on other measures, suggesting it is not a meaningful effect in terms of understanding, behavioural expectations or uptake expectations.

Demographic factors affected understanding of residual risk. Understanding was lower as education level and numeracy decreased and lower in groups self-classifying as Black and Mixed ethnicity compared to White British. This mirrors findings in other risk communication trials, where higher understanding is associated with higher education^4,27,28^, higher numeracy^27^ and White British ethnicity^27^.

Communicating the need to maintain adherence to protective behaviours following a negative test result did not increase expectations of engaging in protective behaviours, although these may have been subject to ceiling effects given the high reported likelihood of engaging in protective behaviours across the sample (*M*=6.4, *SD*=0.9). This finding is akin to other similar Covid-19 vaccine communications tested during lockdown^22^. Information about post-test result behaviours did increase expectations to follow coronavirus guidelines, although this was a very small effect (η2=.004) whereby both those who did and did not receive information about protective behaviours indicated they would follow guidelines as strictly as before. Participants who believe there to be no residual risk of infectiousness following a negative test result were more likely to report they expect to follow guidelines than those who correctly understood residual risk. This exploratory result is difficult to explain and would warrant replication as a pre-planned hypothesis before discussing further.

### Strengths and weaknesses of the study

This study provides the first experimental evidence that some misunderstand there to be no residual risk of infectiousness following a negative asymptomatic Covid-19 test result, while demonstrating the effectiveness of simple, low-cost interventions to increase understanding. Implementing these interventions would be a valuable step in ensuring that the implications of asymptomatic LFD testing are more often understood by the public.

The study has several limitations. First, participants were responding to a hypothetical test result. The interventions would benefit from being tested in a real world setting to check that the increase in understanding is maintained. Second, expectations of engaging in protective behaviours were high. This could have been due to national lockdown restrictions being in place at the time, as in previous studies^22,29^. As restrictions ease, there might be more variability in the propensity to follow guidelines and more pronounced effects of messaging on behaviour. Third, a quota sample was used. Although it was broadly demographically representative of the UK population, it was limited to internet users and could have been subject to bias^30^. A quota sample was favoured as it enables rapid data collection and can therefore meet the demands of a crisis^31^. Participants were randomly allocated to each message, meaning their effects can be experimentally compared and any issues about representativeness are unlikely to affect the interpretation of the findings.

### Implications for policymakers

The results of this study suggest that adding one sentence to a pre-existing single sentence can increase understanding of the meaning of a negative test result. Including the need to continue engaging in protective behaviours following a negative test result, as in current NHS T&T messaging, may also prevent lowering adherence to guidelines. These findings merit implementation with a nested evaluation to check that the effects observed in this hypothetical study are replicated in a real-world setting. However, stronger messages may be needed in contexts where residual risk of infectiousness is higher than in asymptomatic community testing programmes. Messages which include only negatively framed residual risk information could be more effective than the combined positive and negative framing used in this study^19^.

### Unanswered questions and future research

The effects of education, numeracy and ethnicity on understanding of residual risk were consistent with prior studies on risk communication^4, 27, 28^ and suggest there are additional barriers to understanding in those with low education, low numeracy and Black and Mixed ethnicity. Future research should seek to identify and tackle them, to which end co-producing messages with these populations could be a useful approach^32, 33^. Finally, future research should evaluate the effectiveness of the messages that people receive after a positive LFD test result, in terms of encouraging self-isolation or following up with a PCR test. Ensuring people do self-isolate after a test-positive result is important given recent findings that fewer than 50% of symptomatic individuals fully selfisolate^34^.

## Data Availability

The data will be made available upon acceptance of the manuscript for publication from Open Science Framework: https://osf.io/byfz3/

## Acknowledgements

The authors thank the Winton Centre for Risk and Evidence Communication for designing the infographic, Henry Potts for statistical advice and providing comments on the manuscript, and Louise Smith for providing comments on the manuscript.

## Contributors

TMM framed the broad research question. All authors contributed to conceptualising and designing the study. EB and SB completed the data collection and analysis and drafted the manuscript. All authors contributed to, and approved, the final manuscript.

## Ethics approval

The study was reviewed and approved by Public Health England’s Research and Ethics Governance Group (RD432).

## Transparency statement

The lead author affirms that the manuscript is an honest, accurate, and transparent account of the study being reported; that no important aspects of the study have been omitted; and that any discrepancies from the study as originally planned have been explained.

## Dissemination to participants and related patient and public communities

Participants took part in the study anonymously, meaning the authors do not have the necessary details to send participants the results of the study. These findings have been disseminated to relevant stakeholders across government.

## Footnotes

1 Messages are provided by NHS T&T when communicating test results to those who have taken a lateral flow test at a test site or reported their home test result to NHS T&T. The message communicated by NHS T&T after a negative test result includes further information that we did not include in the messages in this study. The NHS T&T wording tested here is the residual risk sentence ‘It’s likely you were not infectious when the test was done’ which follows the statement of the negative test result, as in this study.

2 To ensure meaningful comparisons between genders, participants who reported their gender as ‘non-binary’ (n=1) or ‘prefer not to say’ (n=2) were excluded from the logistic regression analysis given low numbers in each group. When included in the analysis, their understanding of residual risk was not significantly different from the reference category (male) nor did this alter the significance or direction of the other effects or analyses.

## References

1. BBC News. Covid: Tests to be offered twice-weekly to all in England. April 2021. Covid: Tests to be offered twice-weekly to all in England - BBC News [accessed 03 June 2021]

2. García-Fiñana M, Hughes D, Cheyne C, Burnside G, Buchan I, Semple C. Innova Lateral Flow SARS-CoV-2 Antigen test accuracy in Liverpool Pilot: Preliminary Data. November 2020. S0925_Innova_Lateral_Flow_SARS-CoV-2_Antigen_test_accuracy.pdf (publishing.service.gov.uk) [accessed 01 February 2021]

3. Mayers C, Baker K. Impact of false-positives and false-negatives in the UK’s COVID-19 RT-PCR testing programme. June 2020. S0519_Impact_of_false_positives_and_negatives.pdf (publishing.service.gov.uk) [accessed 01 March 2021]

4. Marteau TM, Senior V, Sasieni P. Women’s understanding of a “normal smear test result”: experimental questionnaire based study. BMJ 2001;322:526. doi: 10.1136/bmj.322.7285.526.

5. Michie S, Thompson M, Hankins M. To be reassured or to understand? A dilemma in communicating normal cervical screening results. Brit J Health Psych 2004;9:113–23. doi: 10.1348/135910704322778768.

6. Larsen IK, Grotmol T, Almendingen K, Hoff G. Impact of colorectal cancer screening on future lifestyle choices: a three-year randomized controlled trial. Clin Gastroenterol 2007:5;477–483. doi: 10.1016/j.cgh.2006.12.011.

7. Barnett KN, Weller D, Smith S, et al. Understanding of a negative bowel screening result and potential impact on future symptom appraisal and help-seeking behaviour: a focus group study. Health Expect 2017;20:584–92. doi: 10.1111/hex.12484.

8. Barnett KN, Weller D, Smith S, et al. The contribution of a negative colorectal screening test result to symptom appraisal and help-seeking behaviour among patients subsequently diagnosed with an interval colorectal cancer. Health Expect 2018;21:764–73. doi: 10.1111/hex.12672.

9. Ramachandran S, Mishra S, Condie N, Pickles M. How do HIV-negative individuals in subSaharan Africa change their sexual risk behaviour upon learning their serostatus? A systematic review. Sex Transm Infect 2016;92:571–78. doi: 10.1136/sextrans-2015-052354.

10. Pettengill MA, McAdam AJ. Can We Test Our Way Out of the COVID-19 Pandemic? J Clin Microbiol 2020;58:e02225–e12220. doi: 10.1128/JCM.02225-20.

11. Recchia G, Schneider CR, Freeman ALJ. How do the public interpret COVID-19 swab test results? Comparing the impact of official information about results and reliability used in the UK, US and New Zealand: a randomised, controlled trial. MedRxiv 20243840 [Preprint]. December 05, 2020. [cited 2021 Apr 01] https://doi.org/10.1101/2020.12.04.20243840.

12. Galesic M, Garcia-Retamero R, Gigerenzer G. Using icon arrays to communicate medical risks: overcoming low numeracy. Health Psychol 2009;28:210–16. doi: 10.1037/a0014474.

13. Spiegelhalter D, Pearson M, Short I. Visualizing uncertainty about the future. Science 2011;333:1393–1400. doi: 10.1126/science.1191181.

14. The British Psychological Society. Delivering effective public health campaigns during Covid-19. Delivering effective public health campaigns during Covid-19.pdf (bps.org.uk) November 2020 [accessed 03 June 2021]

15. Michie S, Van Stralen MM, West R. The behaviour change wheel: a new method for characterising and designing behaviour change interventions. Implementation science. 2011;6:1–2.

16. Rogers RW. A protection motivation theory of fear appeals and attitude change1. The journal of psychology. 1975;91:93–114.

17. Gantiva C, Jimenez-Leal W, Urriago-Rayo J. Framing messages to deal with the COVID-19 crisis: the role of loss/gain frames and content. Front Psychol 2021;12:29. doi: 10.3389/fpsyg.2021.568212.

18. Jasper JD, Goel R, Einarson A, Gallo M, Koren G. Effects of framing on teratogenic risk perception in pregnant women. The Lancet 2001;358:1237–8. doi: 10.1016/s0140-6736(01)06353-x.

19. Bigman CA, Cappella JN, Hornik RC. Effective or ineffective: Attribute framing and the human papillomavirus (HPV) vaccine. Patient education and counseling 2010;81:S70–6.

20. NHS. Negative test result for coronavirus (COVID-19). March 2021. Negative test result for coronavirus (COVID-19) -NHS (www.nhs.uk) [accessed 01 March 2021]

21. UK Government. (COVID-19) Coronavirus restrictions: what you can and cannot do. March 2021. (COVID-19) Coronavirus restrictions: what you can and cannot do - GOV.UK (www.gov.uk) [accessed 01 March 2021]

22. Kerr JR, Freeman ALJ, Marteau TM, van der Linden S. Effect of information about COVID-19 vaccine effectiveness and side effects on behavioural intentions: two online experiments. Vaccines 2021;9;379. doi: 10.3390/vaccines9040379

23. YouGov. YouGov / Sky Survey Results. December 2020. Survey Report (yougov.com) [accessed 01 March 2021]

24. Galesic M, Garcia-Retamero R. Statistical numeracy for health: a cross-cultural comparison with probabilistic national samples. Arch Intern Med 2010;170:462–8. doi: 10.1001/archinternmed.2009.481

25. UK Government. Liverpool Covid-19 community testing pilot: interim evaluation report summary.mJanuary 2021. Liverpool COVID-19 community testing pilot: interim evaluation report summary - GOV.UK (www.gov.uk) [Accessed 14 July 2021]

26. Weisberg DS, Keil FC, Goodstein J, Rawson E, Gray JR. The seductive allure of neuroscience explanations. Journal of cognitive neuroscience. 2008;20:470–7.

27. Tait AR, Zikmund-Fisher BJ, Fagerlin A, Voepel-Lewis T. Effect of various risk/benefit trade-offs on parents’ understanding of a pediatric research study. Pediatrics 2010;125(6):e1475–82. doi: https://doi.org/10.1542/peds.2009-1796

28. Treschan TA, Scheck T, Kober A, et al. The influence of protocol pain and risk on patients’ willingness to consent for clinical studies: a randomized trial. Anesthesia & Analgesia 2003;96(2):498–506. doi: 10.1213/00000539-200302000-00037

29. Waller J, Rubin GJ, Potts HWW, Mottershaw AL, Marteau TM. ‘Immunity Passports’ for SARS-CoV-2: an online experimental study of the impact of antibody test terminology on perceived risk and behaviour. BMJ Open 2020;10. doi: 10.1136/bmjopen-2020-040448

30. Office for National Statistics. Internet users, UK: 2019. May 2019. Internet users, UK – Office for National Statistics (ons.gov.uk) [Accessed 14 July 2021]

31. Rubin GJ, Amlôt R, Page L, Wessely S. Methodological challenges in assessing general population reactions in the immediate aftermath of a terrorist attack. Int J Methods Psychiatr Res 2008;17:S29–S35.

32. Li H. Communication for coproduction: Increasing information credibility to fight the coronavirus. The American Review of Public Administration. 2020;50(6-7):692–7.

33. Turk E, Durrance-Bagale A, Han E, et al. International experiences with co-production and people centredness offer lessons for covid-19 responses. BMJ 2021;372:m4752.

34. Smith LE, Potts HWW, Amlot R, Fear NT, Michie S, Rubin GJ. Adherence to the test, trace, and isolate system in the UK: results from 37 nationally representative surveys. BMJ 2021;372:n608. doi: 10.1136/bmj.n608.

