## Supplementary File for "Reducing false reassurance following negative results from asymptomatic coronavirus (Covid-19) testing: an online experiment"

Supplementary material

**Instructions**

You will be asked to imagine that you have participated in a mass testing programme for coronavirus. You will be presented with test results and answer a series of questions about this.

Please read the information carefully as afterwards we will ask you some questions about it, including testing if you remember what the information was.

**Scenario**

Imagine that you have agreed to take part in a mass testing programme for coronavirus in your local area. The programme intends to test as many people as possible who are not currently experiencing symptoms using rapid lateral flow tests.  

You arrive at the test site and are tested using a lateral flow test which involves taking a swab from the back of the throat or the nose. You then leave the test site and are told you will be sent results in approximately 30 minutes. 

Half an hour later, you receive your test results.

**Messages**

*Note all messages were displayed with the same font size.*

**Condition 1** – No residual risk information, no behavioural implications

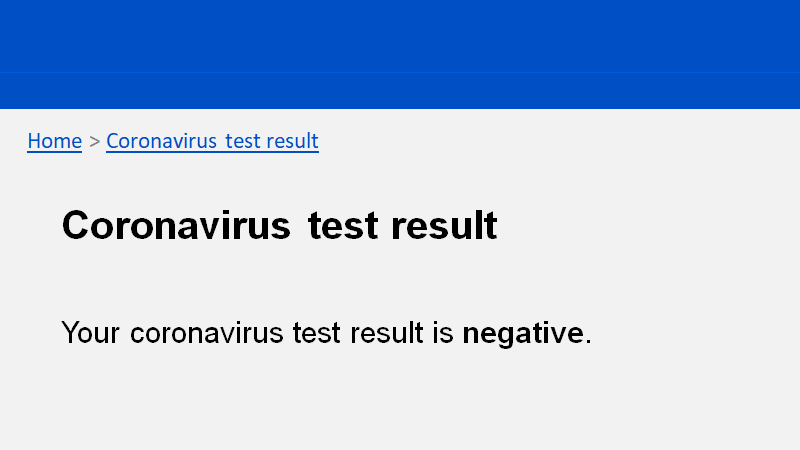

**Condition 2** – Current NHS Test & Trace message, no behavioural implications

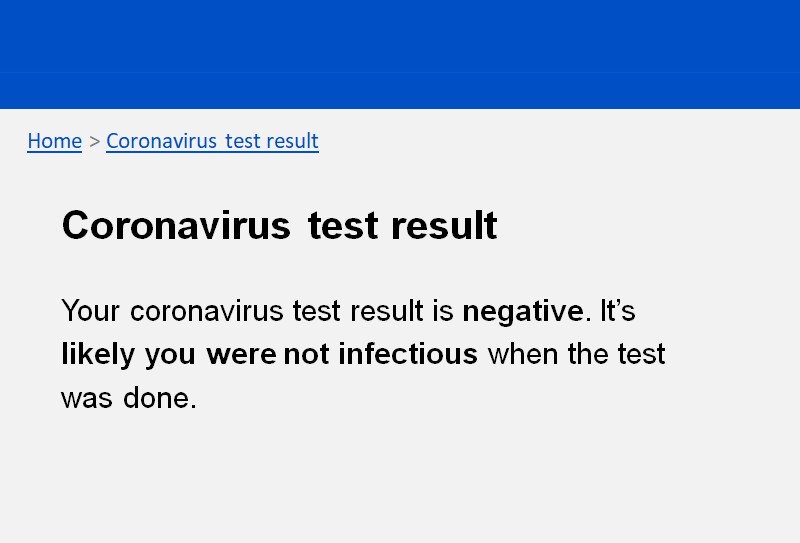

**Condition 3** – Elaborated NHS Test & Trace message, no behavioural implications

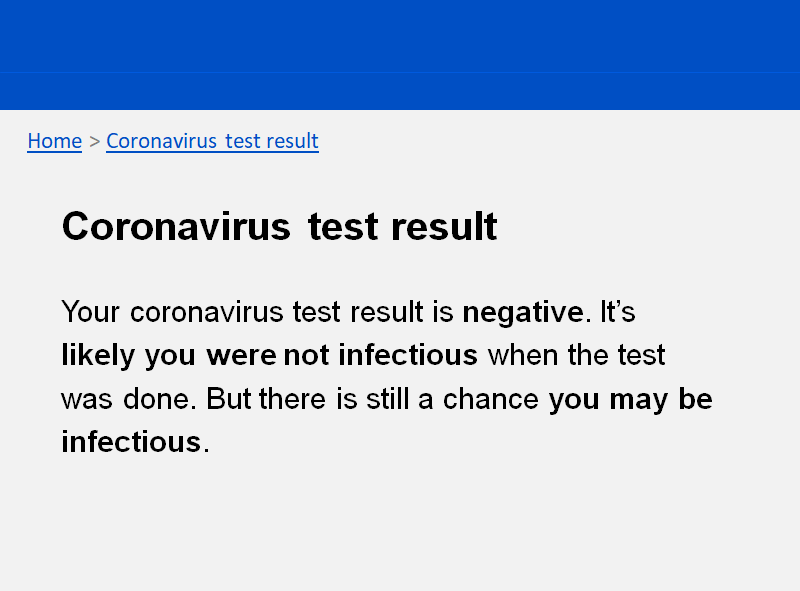

**Condition 4** – Elaborated NHS Test & Trace message with infographic, no behavioural implications

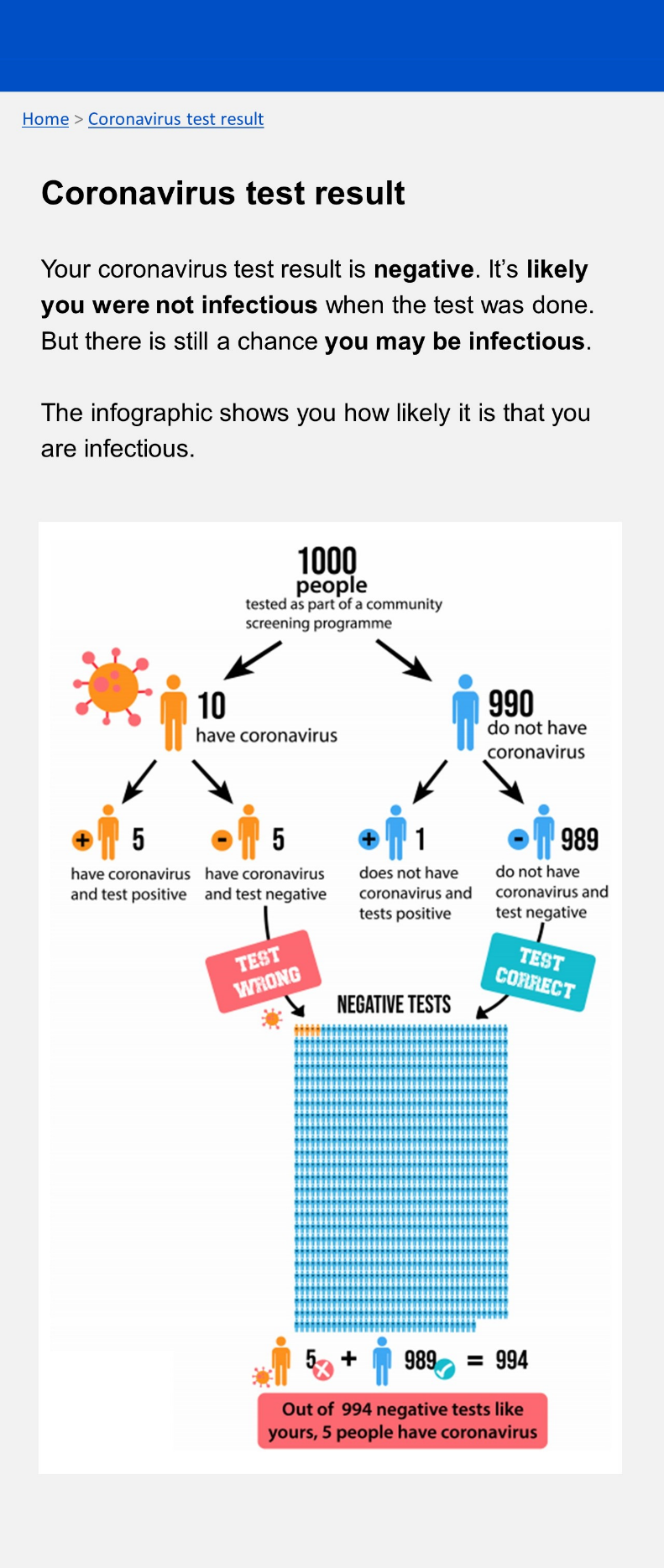

**Condition 5** – No residual risk information, with behavioural implications

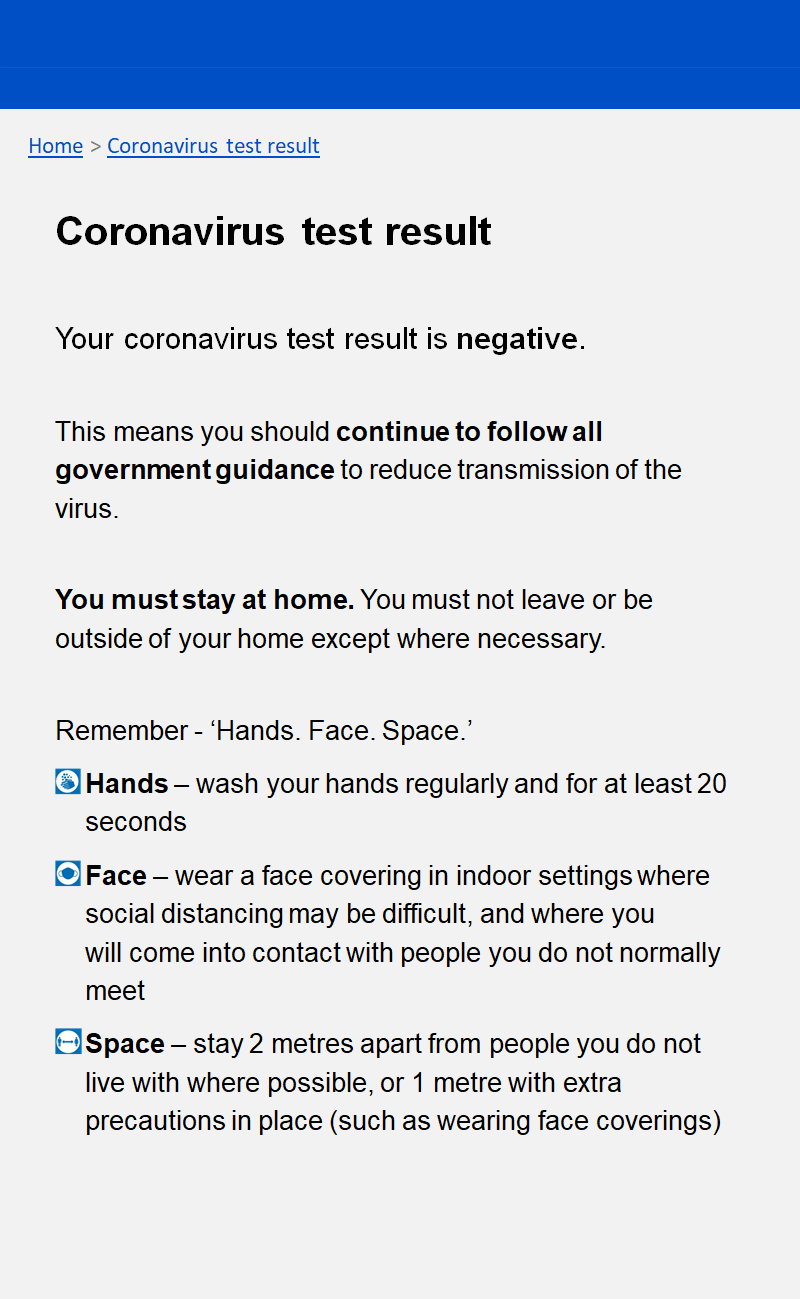

**Condition 6** – Current NHS Test & Trace message, with behavioural implications

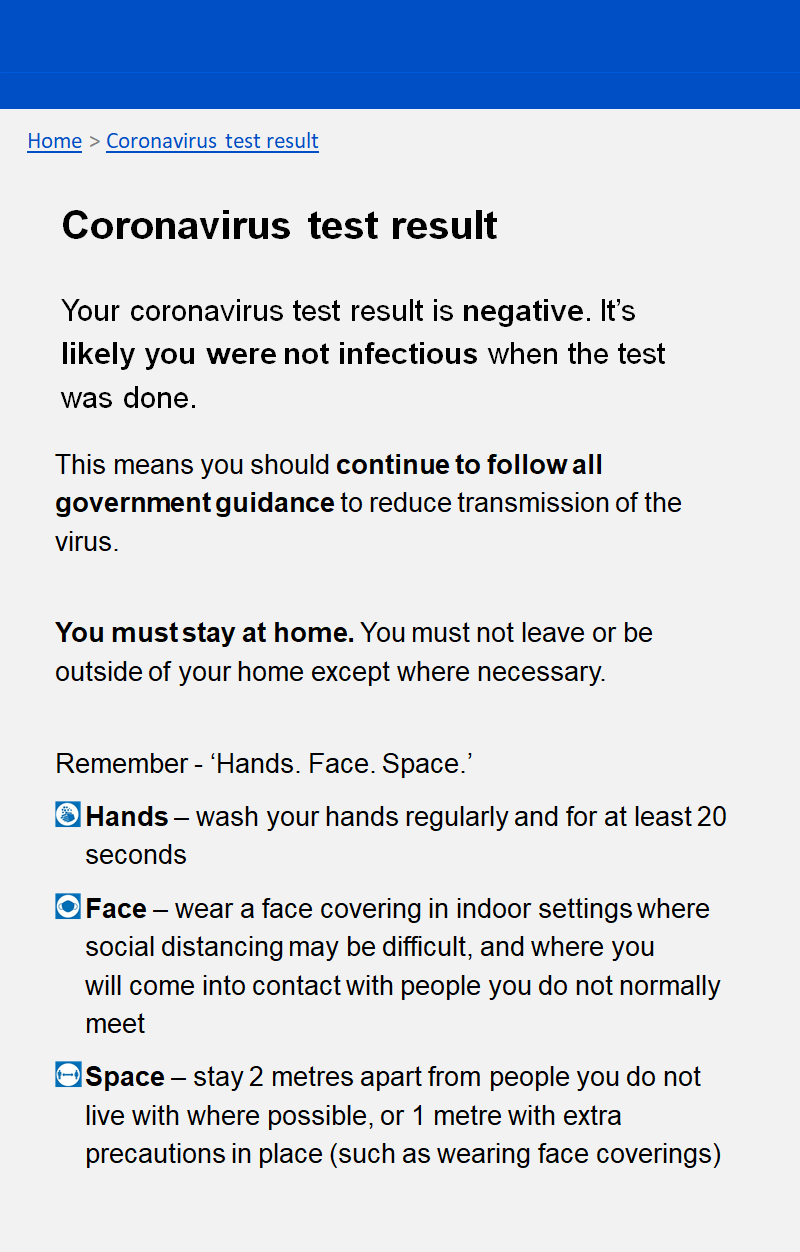

**Condition 7** – Elaborated NHS Test & Trace message, with behavioural implications

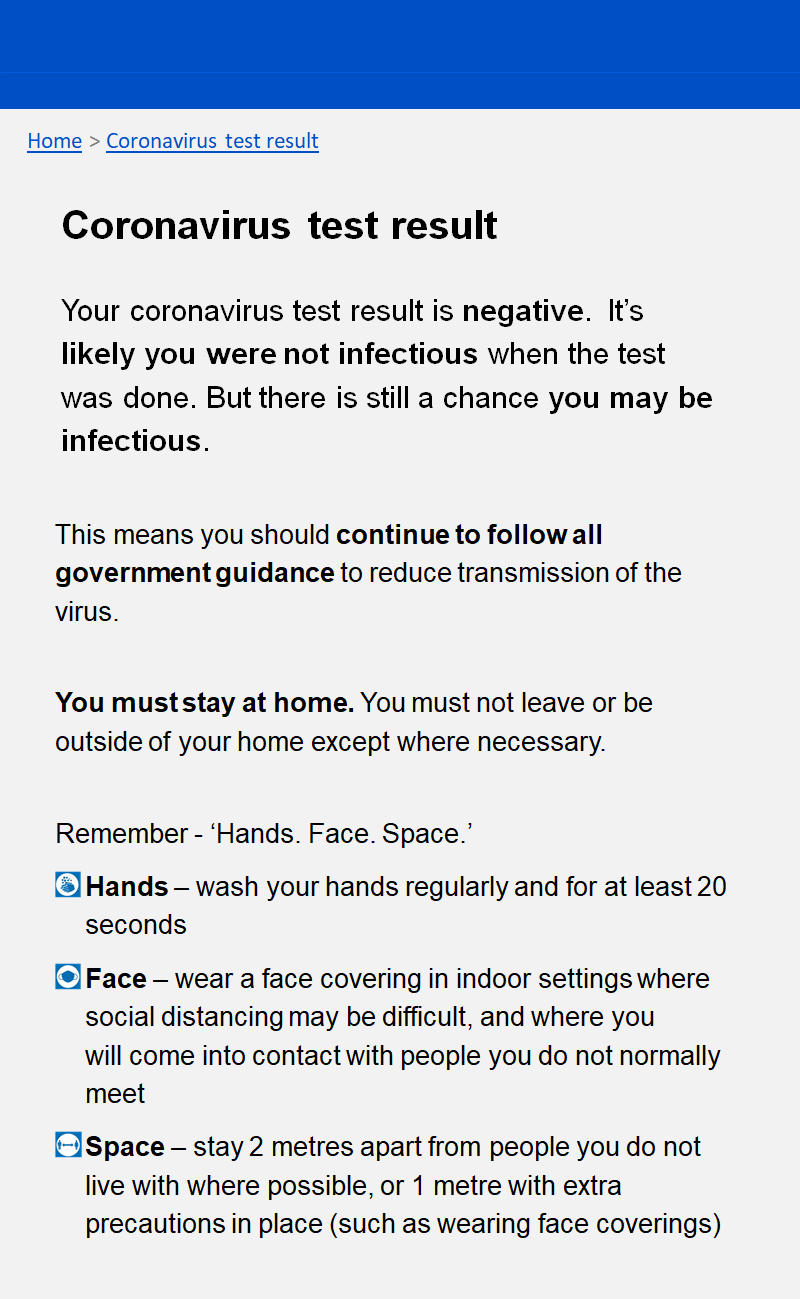

**Condition 8** – Elaborated NHS Test & Trace message with infographic and behavioural implications

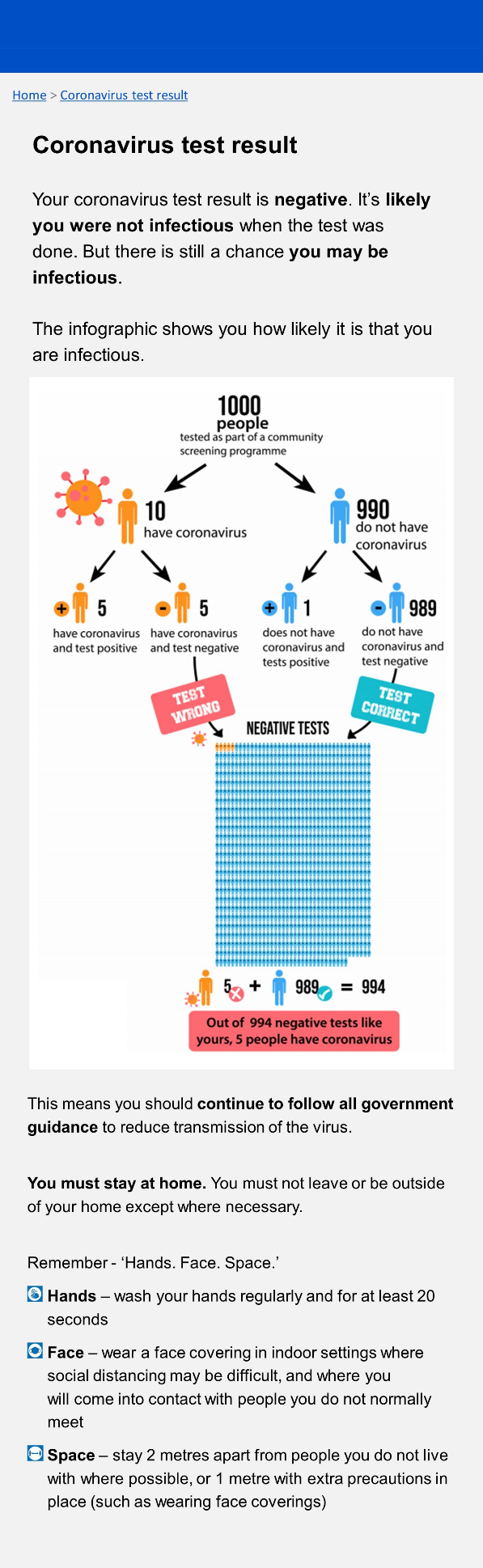

**Question set**

Primary outcome measures and attention check

*Understanding of residual risk*

**Having received this test result, which one of the following statements is true?** 
1. I am not infectious with coronavirus 
2. I am most likely not infectious with coronavirus 
3. I am most likely infectious with coronavirus 
4. I am infectious with coronavirus

*Confidence in understanding*

**How confident are you that you have answered the previous question correctly?**

5- Extremely confident

4- Very confident

3- Moderately confident

2- Slightly confident

1- Not at all confident

*Attention check question*

**How worried would we be if you didn't pay attention? To check that you are paying attention, please do not select an answer below.**

a. Strongly agree
b. Somewhat agree
c. Neither agree nor disagree
d. Somewhat disagree
e. Strongly disagree
f. Don’t know

*Behavioural intention* - general behaviours

**Having received this test result, how strictly would you follow coronavirus guidelines now compared to before taking the test?**

7. A lot more strictly

6. More strictly

5. Slightly more strictly

4. The same as before

3. Slightly less strictly

2. Less strictly

1. A lot less strictly

*Behavioural intention - Specific protective behaviours*

**After receiving this test result, how likely is it that you would engage in the following behaviours because of coronavirus?**

> Social distancing - staying more than 1m from people not in your bubble

> Washing your hands carefully and frequently

> Wearing a face covering in indoor public spaces

> Avoiding meeting with others

> Working from home whenever possible

> Avoiding public transport whenever possible

7 - Very likely

6 - Moderately likely

5 - Slightly likely

4 - Neither likely nor unlikely

3 - Slightly unlikely

2 - Moderately unlikely

1 - Very unlikely

Secondary outcome measures

*Perceived test accuracy*

**How accurate do you think rapid lateral flow tests for coronavirus are?**  (The test you imagined doing in this study was a rapid lateral flow test)

7 - Very accurate

6 - Moderately accurate

5 - Slightly accurate

4 - Neither accurate nor inaccurate

3 - Slightly inaccurate

2 - Moderately inaccurate

1 - Very inaccurate

*Testing uptake intentions*

**If available to you, how likely are you to take a rapid lateral flow test in the future?**

7. Very likely

6. Moderately likely

5. Slightly likely

4. Neither likely nor unlikely

3. Slightly unlikely

2. Moderately unlikely

1. Very unlikely

*Previous testing behaviour*

**When was the last time you took any type of test for coronavirus?**

a. In the last 2 weeks
b. In the last month
c. In the last 3 months
d. In the last year
e. Never

If answer is a/b/c/d:

**What type of test was the one you took most recently?**

a. Lateral Flow Test (LFT) - commonly used for individuals who are asymptomatic and provides results in approximately 30 minutes
b. Polymerase Chain Reaction (PCR) test – commonly booked through the NHS website and used to test individuals who are showing symptoms. Results take between 1-3 days.
c. Other
d. I don’t know what type of test it was

Numeracy and recognition questions

*Numeracy question*

**Which of the following numbers represents the biggest risk of getting a disease?**

a. 1 in 100
b. 1 in 1000
c. 1 in 10

*Recognition question*

**In this study, what were you told when you received your test result?**

a. Your coronavirus test result is inconclusive
b. Your coronavirus test result is positive
c. Your coronavirus test result is negative

Infographic questions (for those in infographic conditions only)

**To what extent did you find the infographic (the diagram of what a negative test result means) easy or difficult to understand?**

5 - Very easy

4 - Somewhat easy

3 - Neither easy nor difficult

2 - Somewhat difficult

1 - Very difficult

**Do you have any suggestions for how the infographic could be improved?** Text box

Demographic questions

**What is your gender?** Male/Female/Non-binary/Prefer not to say/Other

**How old are you?**  Text box (restricted to numbers between 18 and 100)

**What is your ethnicity?** White British/White other/Asian/Black/Arab/Mixed/Other

**In which part of the UK are you currently based?** Northern Ireland/Scotland/Wales/ England-South East/England-South West/ England – London/ England-East of England/ England – East Midlands/ England West Midlands/ England – North West/ England – North East/ England – Yorkshire and Humber

**What is the highest level of education you have completed?** GCSE or equivalent, A levels or equivalent, undergraduate degree, post graduate master's level, postgraduate PhD level

End of study questions

**Do you have any comments or feedback about the study (e.g. your experience, how it could be improved)?**
